# Gene-Specific Cancer Patterns Among Pathogenic Germline Variant Carriers

**DOI:** 10.64898/2026.01.27.26344970

**Authors:** Gideon Idumah, Daphne Newell, Isabella Ribaudo, Ying Ni, Joshua Arbesman

## Abstract

**Key Points:** *Question:* What are the gene-specific cancer patterns and risks among carriers of pathogenic or likely pathogenic variants in cancer susceptibility genes in a population-based cohort?

*Findings:* In this cohort study of 287,076 participants, carriers of pathogenic or likely pathogenic variants in cancer susceptibility genes had markedly higher cancer prevalence compared to noncarriers (e.g., 80% for MEN1 carriers vs 18.6%), with additional risk observed among monoallelic MUTYH co-carriers. Analyses also identified novel gene-cancer associations.

*Meaning:* These findings highlight substantial heterogeneity in inherited cancer risk and support more individualized strategies for genetic testing, cancer screening, and risk assessment.

**Importance:** We previously reported that >5% of the population carries pathogenic or likely pathogenic variants (P/LPVs) in key cancer susceptibility genes (CSGs). However, gene-specific cancer prevalence, spectrum, burden, lifetime risk, comorbidity, and the risk associated with autosomal recessive (AR) genes among carriers remain incompletely defined.

**Objective:** To investigate cancer prevalence, spectrum, burden, lifetime risk, comorbidity, and the uncover novel associations between cancer susceptibility genes and cancer occurrence in a gene-specific manner.

**Design:** A total of 72 cancer susceptibility genes was analyzed. Cancer diagnoses were identified using SNOMED codes and grouped into 35 categories.

**Setting:** This study utilized short-read whole genome sequencing data and electronic health record (EHR) data from the All of Us controlled-tier database v8.

**Participants:** A total of 287,076 participants with both genomic and EHR data were included in the analysis.

**Main Outcomes and Measures:** The primary outcome was an occurrence of a P/LPV in key CSGs. The other outcome is the incidence of cancer diagnosis. Associations between P/LPVs and overall and site-specific cancer risk were evaluated using regression models adjusted for age, sex, race, and ethnicity.

**Results:** Among genes with ≥10 unique carriers, cancer prevalence was highest for *MEN1* (80%), followed by *TP53* (57.7%), *MLH1* (48.4%), and *MSH2* (47.2%), compared to 18.6% among non-carriers. Carriers of P/LPVs in *BRCA1, BRCA2, MLH1, APC, NF1, PTEN,* and *PALB2* had significantly earlier cancer diagnosis compared to non-carriers. Cancer prevalence was markedly higher in *BRCA1* and *BRCA2* carriers who are also mono-allelic *MUTYH* carriers (75% and 45.5%, respectively) compared with *BRCA1* and *BRCA2* alone (43.2% and 36.5%). Similar enrichment was observed for *CHEK2* and *PALB2* monoallelic *MUTYH* co-carriers. Adjusted survival analysis showed increased cancer risk for *MLH1* (OR=6.08), *PTEN* (OR=5.80), and *MSH2* (OR=5.19). Cancer specific analysis reveals significant novel associations including *MITF* with anal/perianal and prostate cancer; *BLM* with ovarian and soft tissue/sarcoma; *WRN* with gynecologic cancer (NOS); and *FH* with hematologic malignancy, alongside well-established gene-cancer relationships.

**Conclusions and Relevance:** This population-based analysis defines gene-specific cancer prevalence, spectrum, and risk, including contributions from AR variants. These findings support more precise genetic testing, screening, and risk stratification for individuals carrying inherited P/LPVs.

## INTRODUCTION

Although significant progress has been made in understanding germline genetic variants in cancer susceptibility genes, critical questions remain regarding lifetime cancer risk, age at cancer diagnosis, overall cancer burden, prevalence, and the spectrum of cancers among individuals with pathogenic or likely pathogenic variants (P/LPVs). The risk associated with variants in autosomal recessive (AR) genes for carriers is also insufficiently characterized.^1,2^ Recent studies have established strong associations between high-penetrance genes and specific cancers.^3–6^ However, most research has relied on case-control cohorts and referral-based or gene-specific registries with less diverse, high-risk populations. These limitations restrict the generalizability of findings for population-scale estimates of penetrance, heterogeneity, and multimorbidity.

The emergence of electronic health record (EHR)-linked population-based research datasets with genomic data has enabled large-scale investigations of individuals with germline variants. Notably, the All of Us (AoU) Research dataset has greater data scale and depth compared to others such as Penn Medicine Biobank, UK Biobank, and Mass General Brigham Biobank. It has unmatched diversity and representation, and its cloud-based access makes it easily accessible to any researcher. Our recent research using AoU dataset indicates that more than 5% of the population carries P/LPVs in key cancer susceptibility genes.^7^ Additional studies suggest that the prevalence of P/LPVs in the general population is substantially higher than previously estimated.^8,9^

This study utilizes US population-based whole-genome sequencing and the longitudinal EHR-linked AoU dataset to systematically examine cancer burden, cancer spectrum, prevalence, and the associations between cancer susceptibility genes and cancer occurrence in a gene-specific manner. The analysis also provides a comprehensive assessment of comorbidity, AR genes, and lifetime risk.

## METHODS

### Study Design and Population

This analysis utilized the AoU research database v8, comprising 633,547 total participants with 414,830 individuals with short-read whole-genome sequencing (srWGS) data. Phenotypic information was derived from the ‘Conditions’ concept set within the EHR., which includes approximately 32,855 standardized clinical findings in 354,581 participants. Our previous work identified 20,968 participants with P/LPVs in 72 cancer susceptibility genes (CSGs), which form the basis for the present analysis. Unless otherwise specified, all analyses were restricted to the 287,076 individuals with both phenotype and genotype information.

### Cancer categorization

Cancer conditions were identified using SNOMED codes by searching for keywords including “adenoc”, “canc”, “malig”, and “carc” in the standard concept name column of the conditions table. The resulting concept names underwent rigorous accuracy review (J.A., Y.N.) to ensure retention of only true cancer concept names. Similar cancer conditions were then consolidated into unified names using regular expression pattern matching of key terms (supplementary material). The analysis identified 66,380 individuals with at least one cancer diagnosis, of whom 54,574 have srWGS data.

### Autosomal Recessive Genes

Among the 72 CSGs, the autosomal recessive (AR) genes include *MUTYH, BLM, RAD50, NBN, DIS3L2, MSH3,* and *WRN*. Carriers of P/LPVs in these genes are separated into two categories. The first category called bi-allelic AR carriers include individuals who are either carriers of a homozygous variant, compound heterozygotes (heterozygous in multiple SNPs within the same AR gene), or having a Variant Allele Frequency (VAF) of at least 85%. Those not meeting these criteria were classified as mono-allelic AR carriers.

### Statistical Analysis

Analyses and statistical comparisons were conducted in the AoU researcher workbench using Python 3 and R in Jupyter notebooks. The Mann-Whitney U Test was used to compare age between groups. Multivariable logistic regression estimated odds ratios for developing cancer, adjusting for age, sex (Male, Female, Others), race (Black, White, Asian, American Indian/Alaska Native, Middle Eastern/North African, Native Hawaiian/Pacific Islander, and Others), and ethnicity (Hispanic, Non-Hispanic, and Others). Categorical variables were one-hot encoded using OneHotEncoder in the scikit-learn library, with White, Female, and Hispanic as reference categories. The age variable was standardized using StandardScaler. The Cox proportional hazards model, implemented with the lifelines and scikit-survival libraries in Python, estimated hazard ratios and their 95% confidence intervals, adjusting for the same covariates as in the logistic regression model. Genes with fewer than 30 carriers and fewer than 5 cancer cases were excluded from both models. For per-cancer inference, each cancer type was required to have at least 200 individuals and at least 20 CSG carriers. Multiple testing was corrected using the false discovery rate (FDR). A significance threshold of p < 0.05 was applied, unless otherwise specified.

## RESULTS

### Cancer Prevalence and Spectrum Among Carriers of P/LPVs in Non-AR Genes

Among individuals with cancer diagnoses and srWGS data, 3713 (6.8%) were CSG carriers. The highest cancer prevalence among carriers of P/LP non-AR gene variants was observed for *MEN1* (80%), followed by *TP53* (57.6%), and *MLH1* (48.4%), with the lowest observed for *WT1* (10%) (Figure 1A). The prevalence of multiple primary cancer diagnosis was also greatest among *MEN1* carriers (40%), followed by *TP53* (35%), and *MSH2* (33.3%). The prevalence of specific cancer types varied substantially across non-AR gene carriers (Figure 1B). Breast, colorectal, and skin cancer (not otherwise specified; NOS) were the most frequently diagnosed malignancies. Breast cancer was diagnosed in 24.8% of *BRCA1* carriers, 18.8% of *BRCA2* carriers, and 13.5% of *PALB2* carriers. Colorectal cancer predominated among *MLH1* (32.3%), *MSH2* (29.2%), and *APC* (12.5%) carriers. Skin cancer (NOS) was most frequently observed among carriers of *TP53* (19.7%), *MSH2* (18.1%), *POT1* (11.1%), and *CDKN2A* (16.3%).

**Figure 1:**
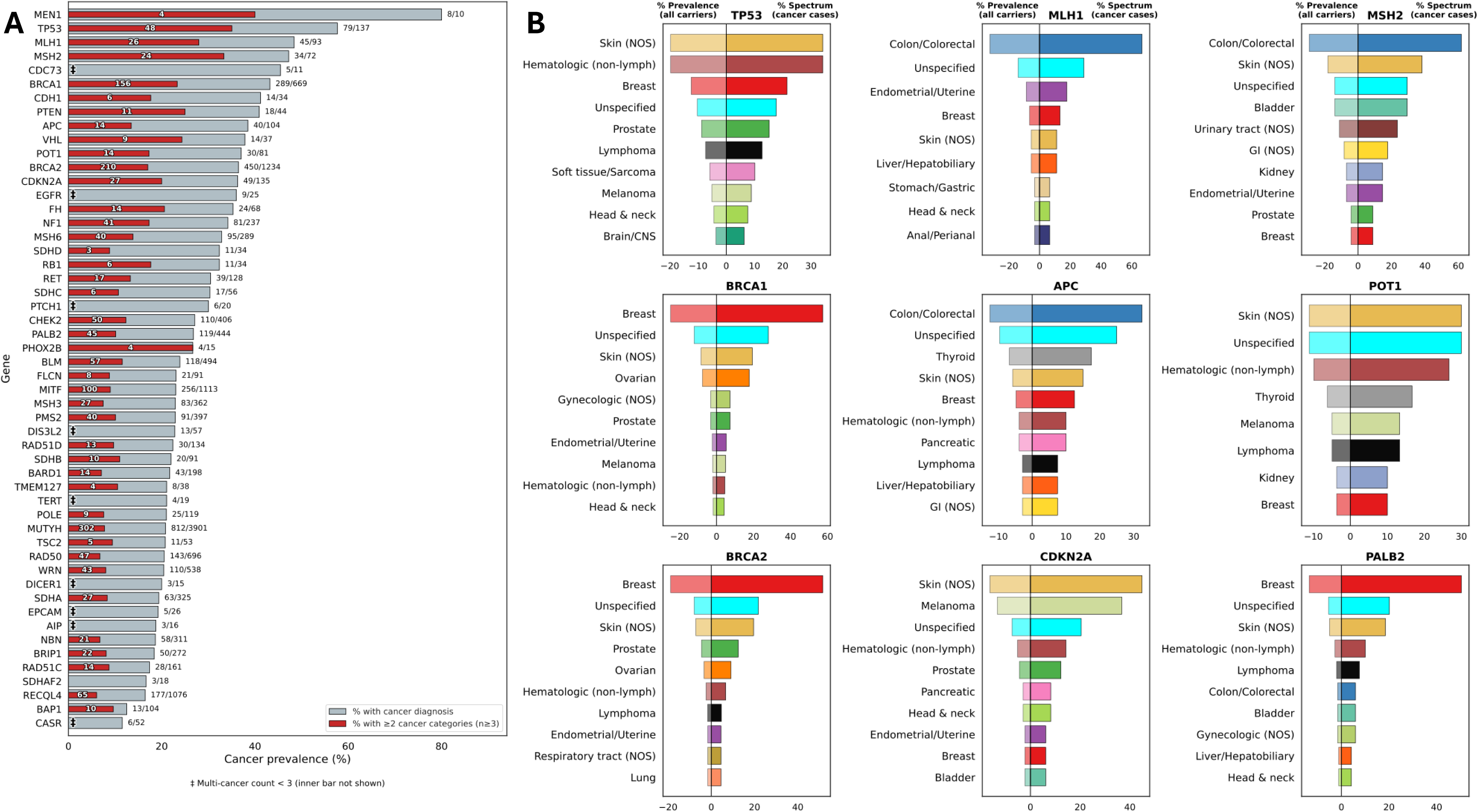
Cancer Prevalence and Spectrum Among Carriers of P/LPVs. *Panel A*: Bar plot showing overall cancer prevalence, and multi-cancer prevalence among individuals with both phenotype and genotype data in cancer susceptibility genes. We did not show genes whose number of carriers is less than 10. *Panel B*: Cancer prevalence (left) and cancer spectrum (right) for top genes illustrated using a butterfly plot. The denominator for prevalence is the number of carriers who have phenotype data for that gene, while the denominator for spectrum is the number of carriers who have cancer.

The overall cancer spectrum also differed by gene (Figure 1B). For example, *POT1* carriers demonstrated a relatively even distribution of cancer types, with each accounting for approximately 10-30% of cancer diagnoses. In contrast, *PALB2* carriers exhibited a more concentrated spectrum; breast cancer comprised approximately 50% of cancer diagnoses, with substantially lower frequencies of other cancers.

### Cancer Prevalence and Spectrum Among Carriers of P/LPVs in AR Genes

Among monoallelic AR gene variant carriers, overall cancer prevalence ranged from 19% for *NBN* to 24% for *BLM* (Figure 2A). For genes with identified biallelic carriers, cancer prevalence was higher among biallelic carriers compared to monoallelic carriers, including *MUTYH* (43% vs 21%) and *WRN* (100% vs 20%).

**Figure 2:**
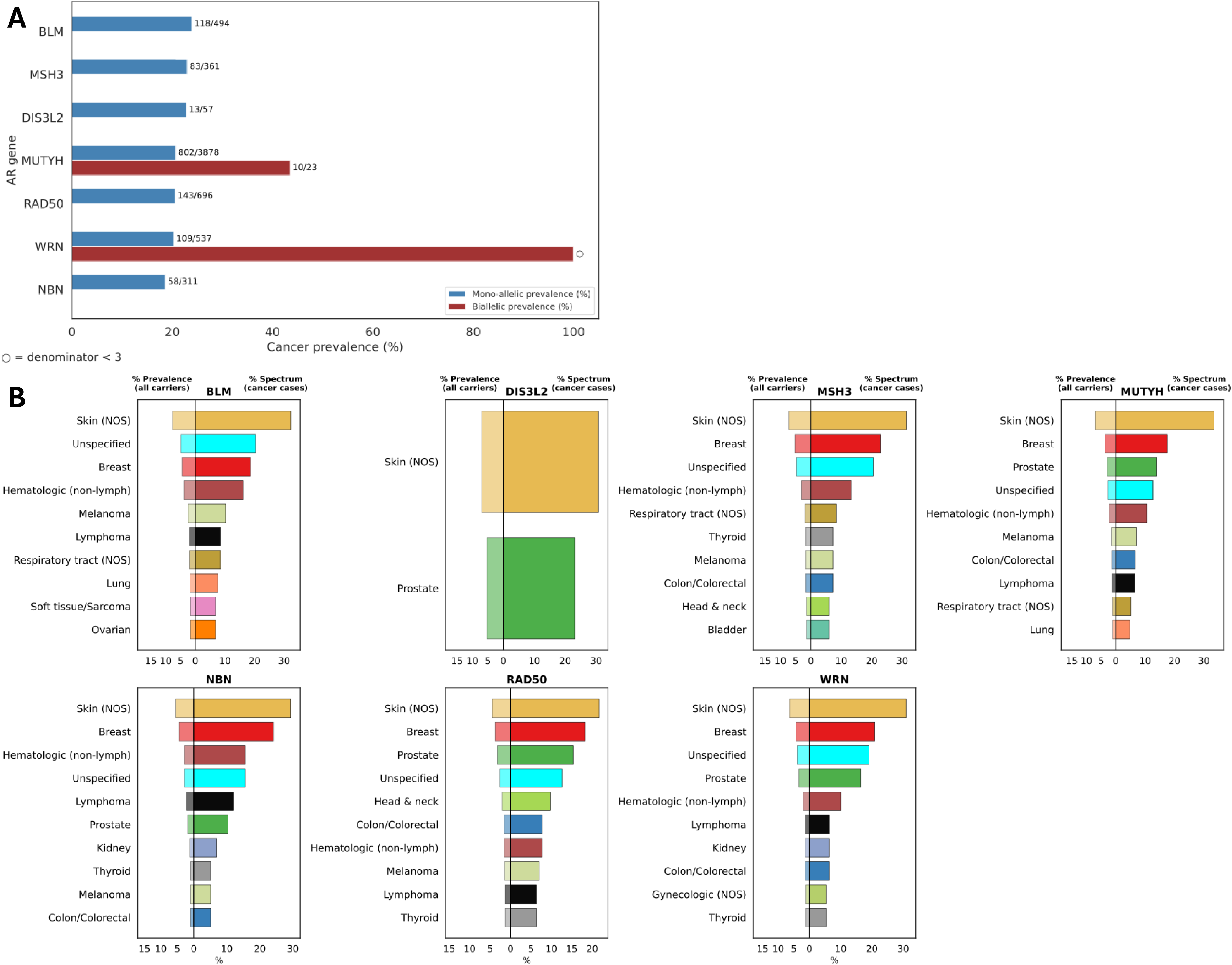
Analysis of Age at first cancer diagnosis for carriers of P/LPVs. *Panel A*: Cancer-free survival probability curves for genes whose carriers have a significantly early onset of cancer compared to non-carrier. *Panel B*: Scatter plot for median age at cancer onset and cancer prevalence across our cancer susceptibility genes, highlighting genes with significantly early onset of cancer.

Cancer type also varied among monoallelic AR gene variant carriers (Figure 2B). Skin cancer (NOS) was the most frequently diagnosed malignancy among AR variant carriers, accounting for approximately 20-30% of cancer diagnoses per gene. Breast, prostate, and hematologic malignancies (excluding lymphoma) were more prevalent than other cancer types. While most AR gene variant carriers demonstrated broad cancer spectra (≥10 cancer types), *DIS3L2* carriers demonstrated a more restricted profile, limited to skin cancer (NOS) and prostate cancer.

### Cancer-Free Survival and Age at Diagnosis

Carriers of non-AR gene variants demonstrated significantly worse cancer-free survival than non-carriers (Figure 3A). Approximately 50% of non-carriers had received a cancer diagnosis by age 78.7. However, among variant carriers, *PTEN* and *MLH1* were associated with the poorest cancer-free survival, with approximately 50% of carriers diagnosed with cancer by age 57.5, compared with age 67 for *BRCA2* and *NF1* carriers and age 73.9 for *PALB2* carriers.

**Figure 3:**
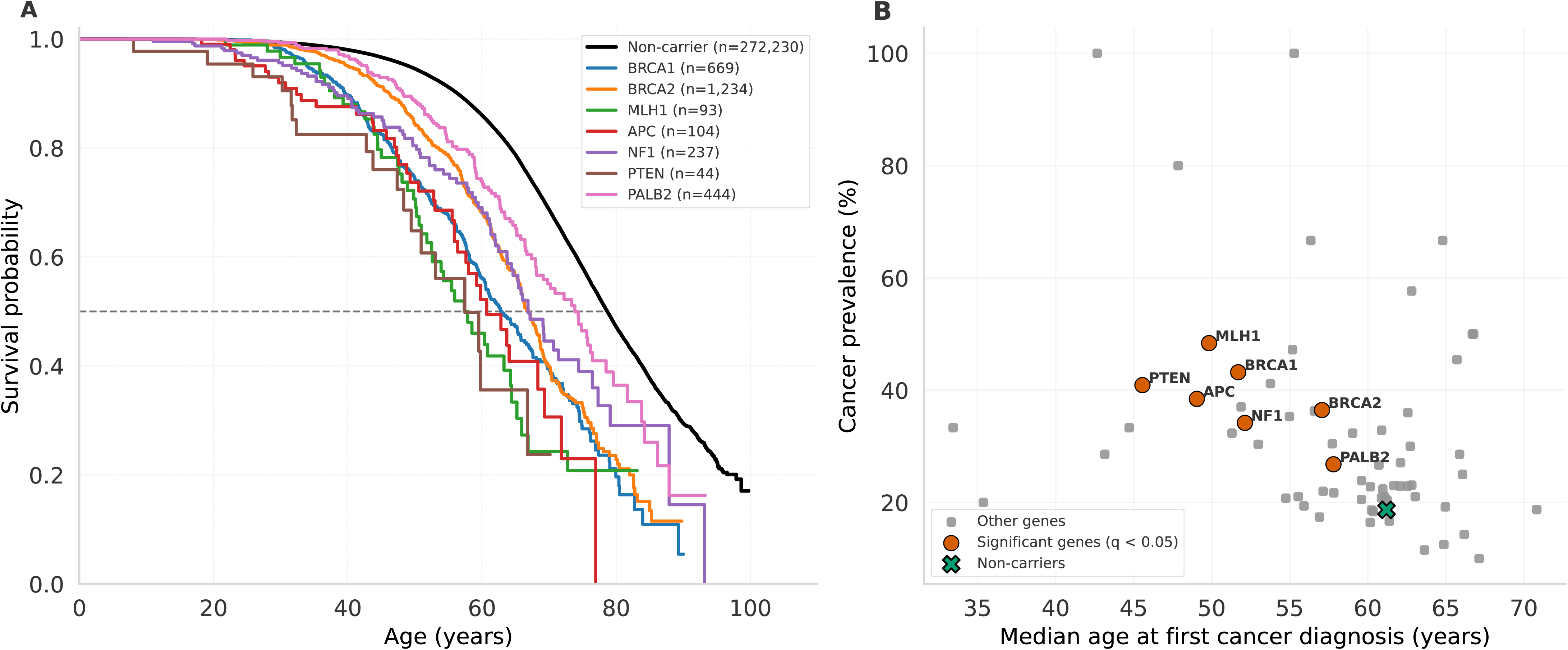
Cancer Prevalence and Spectrum Among Carriers of P/LPVs in Autosomal Recessive Genes. *Panel A*: Cancer prevalence among mono-allelic and bi-allelic autosomal recessive (AR) gene carriers. *Panel B*: Cancer prevalence (left) and cancer spectrum (right) for mono-allelic AR gene carriers illustrated using a butterfly plot. The denominator for prevalence is the number of mono-allelic AR gene carriers who have phenotype data, while the denominator for spectrum is the number of mono-allelic carriers who have cancer.

Additionally, several non-AR gene carriers demonstrated a significantly earlier age at first cancer diagnosis compared to non-carriers, including *BRCA1*, *BRCA2*, *MLH1*, *APC*, *NF1*, *PTEN*, and *PALB2* (Supplementary Figure 1A). The median age at first cancer diagnosis was approximately 61.2 years among non-carriers, compared to 45.6 years for *PTEN*, 51.7 years for *BRCA1*, and 57.8 years for *PALB2*. Across these genes, earlier median age at diagnosis corresponded to a higher overall cancer prevalence (Figure 3B); for example, *PTEN* had a cancer prevalence of approximately 40%, compared to 25% for *PALB2* carriers.

### Co-occurrence of P/LPVs and Associated Cancer Prevalence

We examined the co-occurrence patterns of variants in other cancer susceptibility genes among those who also harbored mono-allelic AR variants (Figure 4A). Among individuals carrying monoallelic AR variants, *MUTYH* exhibited the highest frequency of co-occurring P/LPVs in additional genes. The most common combination was *MUTYH*+*BRCA2*, followed by *MUTYH*+*RECQL4 and MUTYH*+*MITF* (Figure 4A).

**Figure 4:**
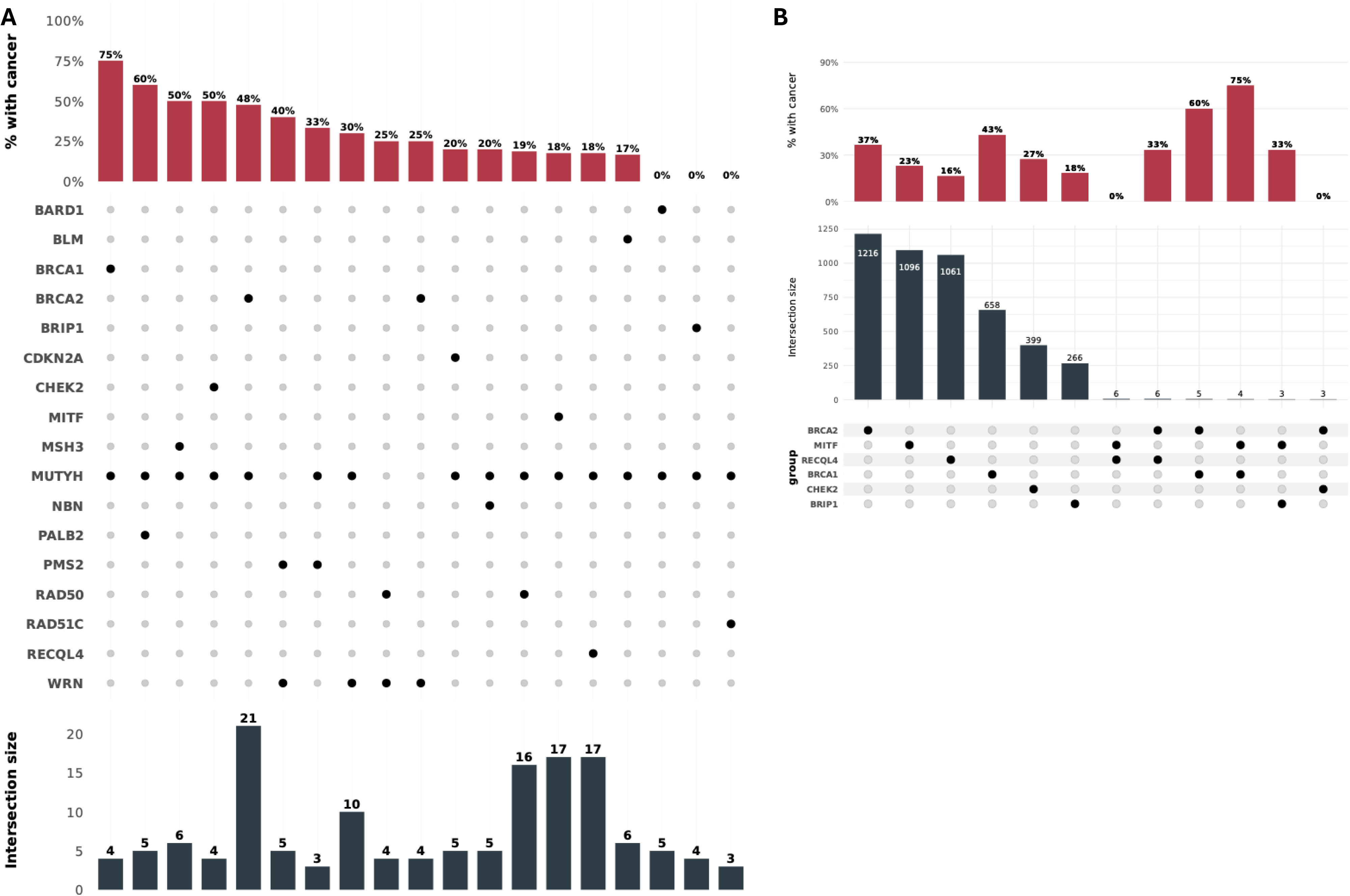
Co-occurrence count and Cancer Prevalence Among Carriers of Multiple P/LPVs. *Panel A*: Upset plot showing the co-occurrence count and cancer prevalence between mono-allelic AR gene carriers that have P/LP variant in other autosomal dominant genes. We do not show counts that are less than 3. *Panel B*: Cancer prevalence and count among co-occurring autosomal dominant (AD) gene carriers. It should be noted that there was no observed co-occurrence between bi-allelic AR carriers and any other genes.

Further analysis of the monoallelic AR carriers with concurrent variants in other genes identified markedly elevated cancer prevalence in select high-risk genotype combinations (Figure 4A). The highest cancer prevalence was observed among individuals with co-occurring *MUTYH*+*BRCA1* variants (75%), followed by those with *MUTYH*+*PALB2* variants (60%), and *MUTYH*+*CHEK2* (50%) (Figure 4A).

Co-occurrence patterns among non-AR genes were also examined (Figure 4B). The most frequent pairings included *RECQL4*+*BRCA2* and *RECQL4*+*MITF*, followed by *BRCA1*+*BRCA2* and *BRCA1*+*MITF*. Increased cancer prevalence was observed among individuals harboring co-occurring variants compared to individuals with only one variant. For example, 75% of co-occurring *BRCA1*+*MITF* carriers have cancer diagnoses, compared with 43% of *BRCA1* carriers and 23% of *MITF* carriers. Similarly, individuals with concurrent *BRCA1*+*BRCA2* variants demonstrated a cancer prevalence of 60%, compared with 36.5% for *BRCA2* carriers alone.

A higher cancer burden was observed in multi-gene carriers compared to single gene carriers (Supplementary Figure 2). Among multi-gene carriers, those with co-occurring *MUTYH*+*WRN* variants had more distinct cancer diagnoses compared to those with other co-occurring variants (Supplementary Figure 2A).

### Multimodal Analysis of Cancer Prevalence and Adjusted Risk Across Germline Genes

Carriers of variants in *MLH1* had the highest adjusted odds of cancer diagnosis compared to noncarriers (adjusted odds ratio [aOR] = 6.08), followed by *PTEN*, *MSH2*, *APC*, and *BRCA1* (aORs = 5.8, 5.19, 4.21, and 4.16, respectively) (Figure 5A). Increasing age was also significantly associated with higher cancer risk (aOR = 2.59). Unknown ethnicity and male sex were also significantly associated with cancer risk, although the magnitude of these effects was modest (aORs = 1.10 and 1.03, respectively). In contract, reduced odds of cancer were observed among participants who self-identified as Hispanic or Latino ethnicity, selected “other” sex, or reported Middle Eastern/North African, other, Asian, Black, or American Indian/Alaskan Native race (aORs = 0.93, 0.89, 0.81, 0.75, 0.62, 0.6, and 0.6, respectively).

**Figure 5:**
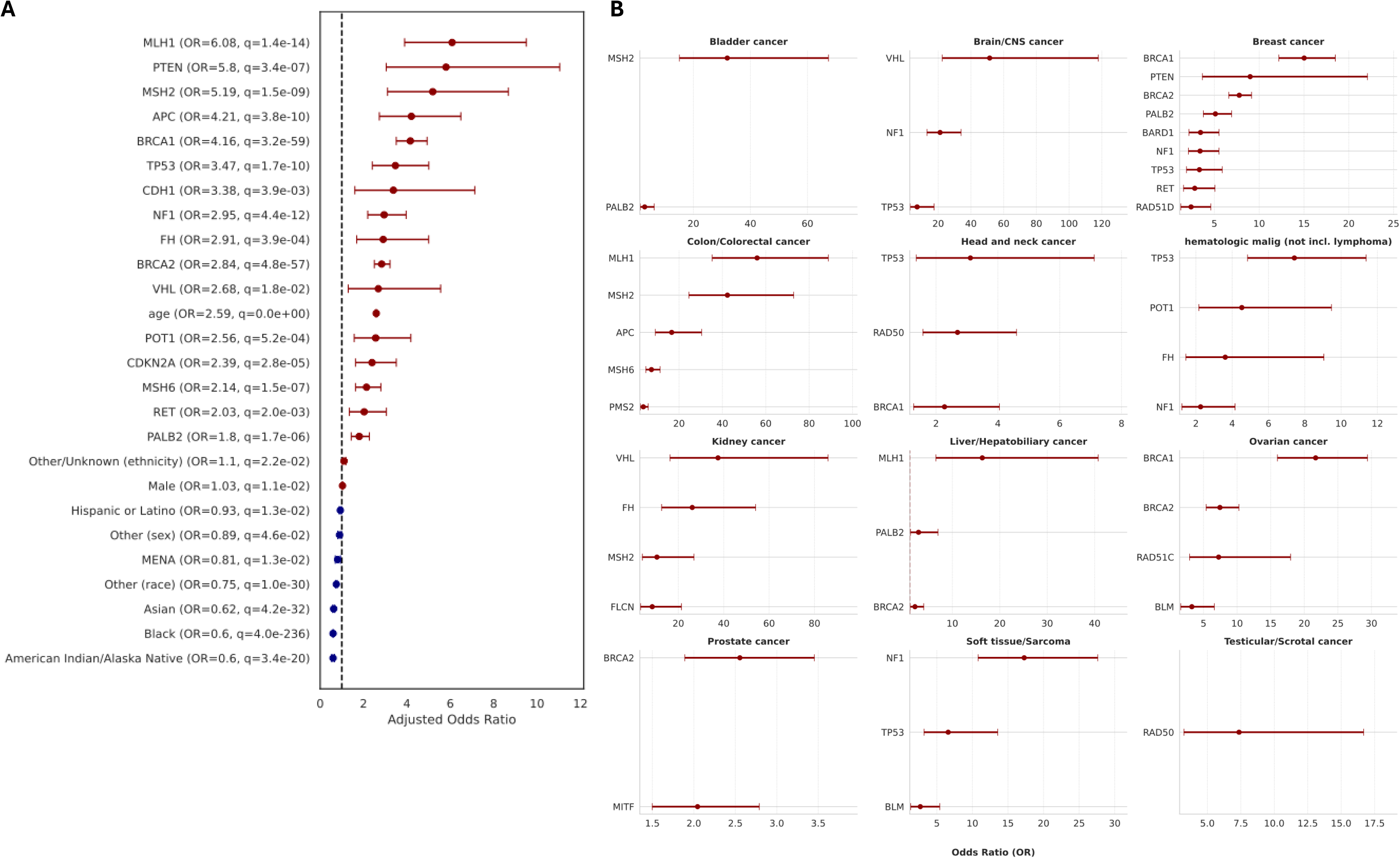
Logistic Regression Inference Result. *Panel A*: Forest plot showing significant features from logistic regression adjusted for age, sex, race, and ethnicity, ranked by adjusted odds ratios. *Panel B*: Per-Cancer forest plots for selected cancer types showing significant genes from logistic regression adjusted for age, sex, race, and ethnicity.

Gene-wise cancer associations across major cancer types were examined with validation of previous associations and identification of novel relationships (Figure 5B). The greatest associations were observed for *MSH2* and urinary tract cancer (NOS), *MLH1* and colon/colorectal cancer, and *VHL* and brain/CNS cancer (ORs = 84.43, 56.01, 51.46). The number of associated genes varied by cancer type; some cancers were associated with only one gene (e.g., lung cancer with *BRCA2* [OR = 2.04], testicular/scrotal cancer with *RAD50* [OR = 7.37], and lymphoma with *TP53* [OR = 4.23]), while others were associated with ≥5 genes, including breast cancer and colon/colorectal cancer. The gene-wise cancer associations with other cancer types are given in supplementary figure 5.

Across genes, we observed a strong positive correlation between cancer prevalence among carriers and the adjusted odds ratio for cancer diagnosis relative to non-carriers (r = 0.93, Supplementary Figure 3B), demonstrating a high concordance between prevalence-based and model-based risk estimates.

Using per-gene Cox proportional hazards models, several genes were significantly associated with earlier time to cancer diagnosis (Supplementary Figure 4B). Carriers of variants in *MSH2* or *TP53* demonstrated the strongest associations with increased cancer hazard (hazard ratios [HRs] = 3.3 and 3.2, respectively). Additional genes significantly associated with increased cancer hazard include *BRCA1*, *MLH1*, *PTEN*, *APC*, and *BRCA2* (HRs = 2.9, 2.9, 2.6, 2.4, and 2.2, respectively).

## DISCUSSION

In this large population-based cohort, we present a gene-specific assessment of cancer prevalence, spectrum, and burden among carriers of P/LPVs across 72 cancer susceptibility genes. Using linked genomic and EHR data from AoU, we identify heterogeneity in cancer risk by gene, including differences in overall prevalence, incidence of multiple primary malignancies, and cancer type distribution. To our knowledge, gene-specific cancer prevalence and patterns across a broad set of cancer susceptibility genes have not been systematically evaluated in a general, unselected population. Analysis within an unselected population allows estimation of cancer risk among variant carriers identified independent of personal or family cancer history, providing clinically relevant context for contrasting risk estimates derived from specialty clinic–based cohorts. Of the 14,819 CSG carriers with EHR information, 3,713 (25%) have a documented cancer diagnosis; the remaining 75% represent an at-risk population in whom appropriate cancer screening and surveillance have substantial potential to reduce morbidity and save lives. The age distribution indicates that carriers without a cancer diagnosis have a significantly lower median age compared to those with documented cancer diagnoses, consistent with increasing cancer risk over time (Supplementary Figure 1B).

Our study found heterogeneity in both cancer prevalence and cancer type across genes, underscoring substantial gene-specific differences in cancer risk. For example, carriers of high-penetrance genes such as *MEN1* and *TP53* showed substantial cumulative cancer burden and multiple primary malignancies, supporting a multisystem surveillance, whereas other genes conferred more moderate risk that may warrant tailored screening approaches. Gene-specific differences in cancer spectrum were evident for *BRCA1* and *BRCA2* despite similar overall cancer prevalence. Breast cancer predominated for both genes and was more frequent among *BRCA1;* ovarian cancer was more common with *BRCA1* and prostate cancer with *BRCA2*, with skin cancer emerging as the second most common malignancy for both. These findings are consistent with prior estimates showing comparable lifetime breast cancer risk for *BRCA1* and *BRCA2* but substantially higher ovarian cancer risk for *BRCA1*.^10,11^ More broadly, skin cancer was among the most frequent malignancies across multiple cancer susceptibility genes, including *CDKN2A, TP53, MSH2, POT1, BRCA1, BRCA2,* and *PALB2*, and was the most prevalent cancer among AR gene carriers. Together with prior evidence demonstrating P/LPVs in approximately 15% of melanoma cases and an increased burden of P/LPVs among individuals with frequent basal cell carcinoma, these findings indicate that inherited susceptibility to skin cancer is more relevant than previously appreciated.^12,13^

Our findings provide additional evidence validating several known gene-cancer associations, including *BRCA1* and *BRCA2* with breast cancer and ovarian cancer,^4^ *PTEN* with thyroid cancer,^14^ *CDKN2A* with melanoma,^12^ and *VHL* and *FH* with kidney cancer.^15,16^ In addition, we identified several novel associations that extend the recognized tumor spectrum of established cancer susceptibility genes. We found that pathogenic *MITF* variants were associated with anal/perianal and prostate cancers, expanding beyond its well-described role in melanoma predisposition.^17^ *BLM* variants were associated with increased risk of ovarian and soft tissue or sarcoma malignancies beyond the canonical Bloom syndrome phenotype.^18^ We identified a novel association between *WRN* and gynecologic cancers not previously described in Werner syndrome–associated cancer risk.^19^ Similarly, the association between *FH* and hematologic malignancies contrasts with existing literature, which predominantly restricts FH-related cancer predisposition to solid tumors.^16^ Consistent with Lynch syndrome, *MLH1* variants were associated with colon/colorectal and endometrial/uterine cancers;^20^ however, the observed association with liver cancer is not well established in existing literature. While these findings warrant cautious interpretation and further validation, collectively, these observations highlight the value of large population-based datasets in uncovering novel gene-specific cancer associations that may help tailor future research.

These findings also draw attention to the role of inheritance patterns in shaping cancer risk. Cancer risk associated with AR cancer susceptibility genes remains less well defined, particularly among monoallelic carriers, and poses ongoing challenges for clinical counseling. In this population-based analysis, cancer prevalence varied by gene, with the highest burden observed among biallelic carriers, as expected. Such patterns are exemplified by classic AR syndromes, including Fanconi anemia, ataxia-telangiectasia, and xeroderma pigmentosum, in which biallelic variants in genes involved in DNA repair and genome stability confer substantial cancer risk.^21,22^ In our cohort, carriers of biallelic *MUTYH* variants demonstrated a high cancer burden consistent with *MUTYH*-associated polyposis; interestingly, monoallelic *MUTYH* carriers also exhibited measurable cancer risk, suggesting that heterozygous carriage may confer modest susceptibility not uniformly addressed in current guidelines.^23^ Similar zygosity-dependent patterns were observed for other AR genes, including *MSH3, NBN,* and *WRN*. When examining overall cancer and multi-cancer prevalence across AR and non-AR genes, we observed that some AR genes demonstrated higher cancer prevalence than non-AR genes. For example, *BLM* mono-allelic variant carriers were found to have an approximately 25% cancer prevalence, surpassing well known non-AR genes such as *BRIP1* and *BAP1*. Together, these results align with emerging evidence that carriers of P/LPVs for AR conditions may also harbor cancer risk.

Beyond single-gene effects, co-occurring P/LPVs were associated with a greater overall cancer burden. Individuals carrying P/LPVs in multiple cancer susceptibility genes demonstrated higher cancer prevalence and greater likelihood of multiple primary malignancies than single-variant carriers, consistent with prior population-based studies showing increasing cancer risk with a greater number of P/LPVs. ^24,25^ This pattern was particularly evident among *BRCA1* and *BRCA2* carriers who also harbored monoallelic *MUTYH* variants, as well as among *CHEK2+PALB2* co-carriers, in whom cancer prevalence exceeded that observed in single-gene carriers. These findings align with existing evidence supporting largely additive or multiplicative effects of co-occurring variants, rather than strong synergistic interactions, with clinical phenotypes generally reflecting the independent cancer spectra of each gene.^26–28^ Together, these results highlight the importance of considering cumulative germline variant burden in risk assessment while recognizing that current evidence supports incremental risk rather than markedly more aggressive disease.^25^

Carriers of P/LPVs developed cancer at younger ages and had shorter cancer-free survival than non-carriers, particularly with high-penetrance genes. *TP53, MEN1,* and *MLH1* carriers showed the earliest ages at diagnosis and the shortest cancer-free survival, consistent with prior reports of early-onset malignancy in Li-Fraumeni syndrome, multiple endocrine neoplasia type 1, and Lynch syndrome.^20,29^ Across genes, earlier age at diagnosis closely tracked with higher cancer prevalence, indicating that early onset reflects greater cumulative risk. The concordance between prevalence estimates, adjusted odds ratios, and hazard ratios supports the clinical robustness of these findings and underscores the importance of gene-specific risk stratification to inform the timing and intensity of cancer surveillance.

## LIMITATIONS

Important limitations include the absence of environmental and lifestyle risk factors, reliance on EHR-derived cancer diagnoses that may be subject to misclassification, and limited statistical power for rare genes and uncommon cancer types. Additionally, demographic categorizations were based on self-identified race and ethnicity in EHRs, which may not fully capture the complexity of genetic ancestry.

## CONCLUSIONS

These population-derived risk estimates refine known gene-cancer associations, uncovers new relations, and supports a shift away from family-based cancer surveillance strategies to gene- and burden-informed approaches. It further demonstrates gene-specific heterogeneity in cancer risk, spectrum, and age at onset, underscoring the value of integrating large-scale genomic data into clinical cancer risk assessment and surveillance frameworks.

## Supporting information

Cancer Mapping - Supplementary File

## Data Availability

All data produced in the present study are available upon reasonable request to the authors

## Acknowledgments

We gratefully acknowledge *All of Us* participants for their contributions, without whom this research would not have been possible. We also thank the National Institutes of Health’s *All of Us* Research Program for making available the participant data examined in this study. This study used data from the *All of Us* Research Program’s Registered Tier Dataset v8, available to authorized users on the Researcher Workbench.

## Author Contribution

Arbesman and Idumah had full access to all the data in the study and take responsibility for the integrity of the data and the accuracy of the data analysis.

*Concept and design*: Idumah, Ni, Arbesman.

*Acquisition, analysis, or interpretation of data*: Idumah, Newell, Ribaudo

*Drafting of the manuscript*: All authors

*Critical revision of the manuscript for important intellectual content*: All authors

*Statistical analysis*: Idumah.

*Supervision*: Ni, Arbesman.

---

**Supplementary Figure 1: Age Analysis Results**

Panel A: Empirical cumulative distribution function result showing the cumulative probability of getting cancer for carriers in genes with significantly early onset of cancer diagnosis compared to non-carriers. Panel B: Age distribution for P/LPVs carriers with and without cancer

**Supplementary Figure 2: Burden Plots for Single and Multi-Carriers of P/LPVs**

Panel A: Burden plot showing the distribution of number of distinct cancer categories for P/LPVs in selected genes using violin plots. Panel B: Burden plot showing the distribution of number of distinct cancer categories for P/LPVs in multiple genes using violin plots.

**Supplementary Figure 3: Result from Cox’s model Analysis**

Panel A: Volcano plot of Cox’s time to event hazard ratio (x-axis) versus the FDR corrected p-values (y-axis) across all genes after adjusting for necessary covariates. Panel B: Relationship between per-gene cancer prevalence and adjusted odds ratio, demonstrating strong internal concordance (r ≈ 0.93).

**Supplementary Figure 4: Forest Plot showing Adjusted Odd and Hazard Ratios for all features**

Panel A: Forest plot showing the odd’s ratio of all features from logistic regression adjusted for age, sex, race, and ethnicity, ranked by adjusted odds ratios. Panel B: Forest plot showing the hazard ratio of all features from logistic regression adjusted for age, sex, race, and ethnicity, ranked by adjusted odds ratios.

**Supplementary Figure 5: Gene-wise Forest Plot for other Cancer Types**

Forest plot showing the association between genes and other cancer types not shown in the main manuscript.

## References

1. Stastna B, Dolezalova T, Matejkova K, et al. Germline pathogenic variants in the MRE11, RAD50, and NBN (MRN) genes in cancer predisposition: A systematic review and meta-analysis. Int J Cancer 2024;155(9):1604–15.

2. Theodoratou E, Campbell H, Tenesa A, et al. A large-scale meta-analysis to refine colorectal cancer risk estimates associated with MUTYH variants. Br J Cancer 2010;103(12):1875–84.

3. Hu C, Hart SN, Gnanaolivu R, et al. A Population-Based Study of Genes Previously Implicated in Breast Cancer. N Engl J Med 2021;384(5):440–51.

4. Breast Cancer Association Consortium. Breast Cancer Risk Genes — Association Analysis in More than 113,000 Women. New England Journal of Medicine 2021;384(5):428–39.

5. Zeng C, Bastarache LA, Tao R, et al. Association of Pathogenic Variants in Hereditary Cancer Genes With Multiple Diseases. JAMA Oncol 2022;8(6):835.

6. Lu H-M, Li S, Black MH, et al. Association of Breast and Ovarian Cancers With Predisposition Genes Identified by Large-Scale Sequencing. JAMA Oncol 2019;5(1):51–7.

7. Idumah G, Newell D, Hadrys M, Ribaudo I, Ni Y, Arbesman J. Pathogenic Germline Variants in Cancer Susceptibility Genes. JAMA 2025;334(19):1765–8.

8. Park J, Karnati H, Rustgi SD, Hur C, Kong X-F, Kastrinos F. Impact of population screening for Lynch syndrome insights from the All of Us data. Nat Commun 2025;16(1):523.

9. White SL, Jamil T, Bell C, et al. Population Prevalence of the Major Thyroid Cancer-Associated Syndromes. J Clin Endocrinol Metab 2025;110(12):e4049–54.

10. Kotsopoulos J, Hathaway CA, Narod SA, et al. Germline Mutations in 12 Genes and Risk of Ovarian Cancer in Three Population-Based Cohorts. Cancer Epidemiol Biomarkers Prev 2023;32(10):1402–10.

11. US Preventive Services Task Force. Risk Assessment, Genetic Counseling, and Genetic Testing for BRCA-Related Cancer: US Preventive Services Task Force Recommendation Statement. JAMA 2019;322(7):652–65.

12. Funchain P, Ni Y, Heald B, et al. Germline cancer susceptibility in individuals with melanoma. Journal of the American Academy of Dermatology 2024;91(2):265–72.

13. Cho HG, Kuo KY, Li S, et al. Frequent basal cell cancer development is a clinical marker for inherited cancer susceptibility. JCI Insight 3(15):e122744.

14. Tan M-H, Mester JL, Ngeow J, Rybicki LA, Orloff MS, Eng C. Lifetime Cancer Risks in Individuals with Germline *PTEN* Mutations. Clinical Cancer Research 2012;18(2):400–7.

15. Shirole NH, Kaelin WG. VHL and HIF at the Center of RCC Biology. Hematol Oncol Clin North Am 2023;37(5):809–25.

16. Lu E, Hatchell KE, Nielsen SM, et al. Fumarate hydratase variant prevalence and manifestations among individuals receiving germline testing. Cancer 2022;128(4):675–84.

17. Roider E, Lakatos AIT, McConnell AM, et al. MITF regulates IDH1, NNT, and a transcriptional program protecting melanoma from reactive oxygen species. Sci Rep 2024;14(1):21527.

18. Hodson C, Low JKK, van Twest S, et al. Mechanism of Bloom syndrome complex assembly required for double Holliday junction dissolution and genome stability. Proceedings of the National Academy of Sciences 2022;119(6):e2109093119.

19. Mukherjee S, Sinha D, Bhattacharya S, Srinivasan K, Abdisalaam S, Asaithamby A. Werner Syndrome Protein and DNA Replication. International Journal of Molecular Sciences 2018;19(11):3442.

20. Bonadona V, Bonaïti B, Olschwang S, et al. Cancer Risks Associated With Germline Mutations in MLH1, MSH2, and MSH6 Genes in Lynch Syndrome. JAMA 2011;305(22):2304–10.

21. Rahman N, Scott RH. Cancer genes associated with phenotypes in monoallelic and biallelic mutation carriers: new lessons from old players. Hum Mol Genet 2007;16(R1):R60–6.

22. Ballinger ML, Goode DL, Ray-Coquard I, et al. Monogenic and polygenic determinants of sarcoma risk: an international genetic study. The Lancet Oncology 2016;17(9):1261–71.

23. Toboeva MK, Хетаговна ТМ, Shelygin YA, et al. MutYH-associated polyposis. Terapevticheskii arkhiv 2019;91(2):97–100.

24. Shevach JW, Xu J, Snyder N, et al. Established Cancer Predisposition Genes in Single and Multiple Cancer Diagnoses. JAMA Oncol 2025;11(10):1222–30.

25. Speicher MR, Geigl JB, Tomlinson IP. Effect of genome-wide association studies, direct-to-consumer genetic testing, and high-speed sequencing technologies on predictive genetic counselling for cancer risk. The Lancet Oncology 2010;11(9):890–8.

26. Pócza T, Papp J, Bozsik A, et al. Double Pathogenic or Likely Pathogenic Variants in Cancer Predisposition Genes in Hungarian Cancer Patients. International Journal of Molecular Sciences 2025;26(17):8390.

27. Infante M, Arranz-Ledo M, Lastra E, et al. Increased Co-Occurrence of Pathogenic Variants in Hereditary Breast and Ovarian Cancer and Lynch Syndromes: A Consequence of Multigene Panel Genetic Testing? International Journal of Molecular Sciences 2022;23(19):11499.

28. Pritchard AL, Johansson PA, Nathan V, et al. Germline mutations in candidate predisposition genes in individuals with cutaneous melanoma and at least two independent additional primary cancers. PLoS ONE 2018;13(4):e0194098.

29. Andrade KC de, Khincha PP, Hatton JN, et al. Cancer incidence, patterns, and genotype–phenotype associations in individuals with pathogenic or likely pathogenic germline TP53 variants: an observational cohort study. The Lancet Oncology 2021;22(12):1787–98.

