## Supplementary figures and images for "Gene-Specific Cancer Patterns Among Pathogenic Germline Variant Carriers"

### Supplemental Figure 1

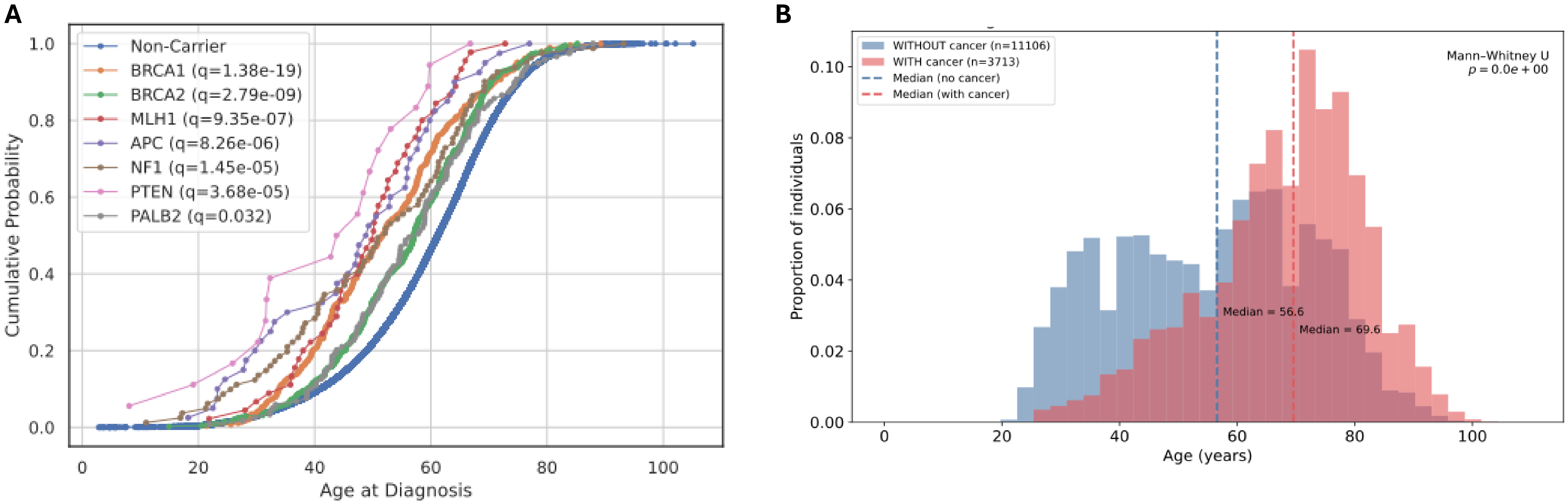

### Supplemental Figure 2

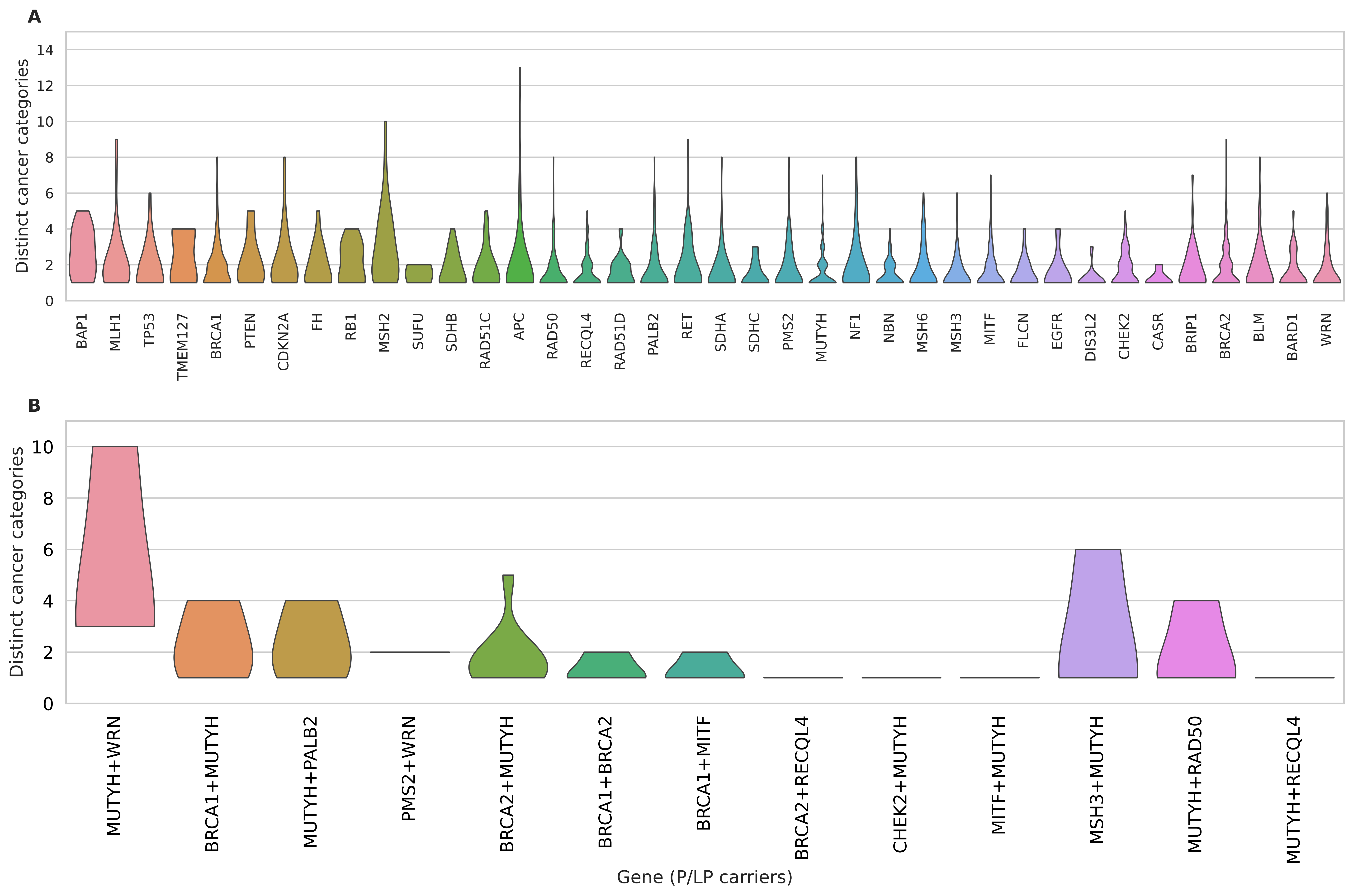

### Supplemental Figure 3

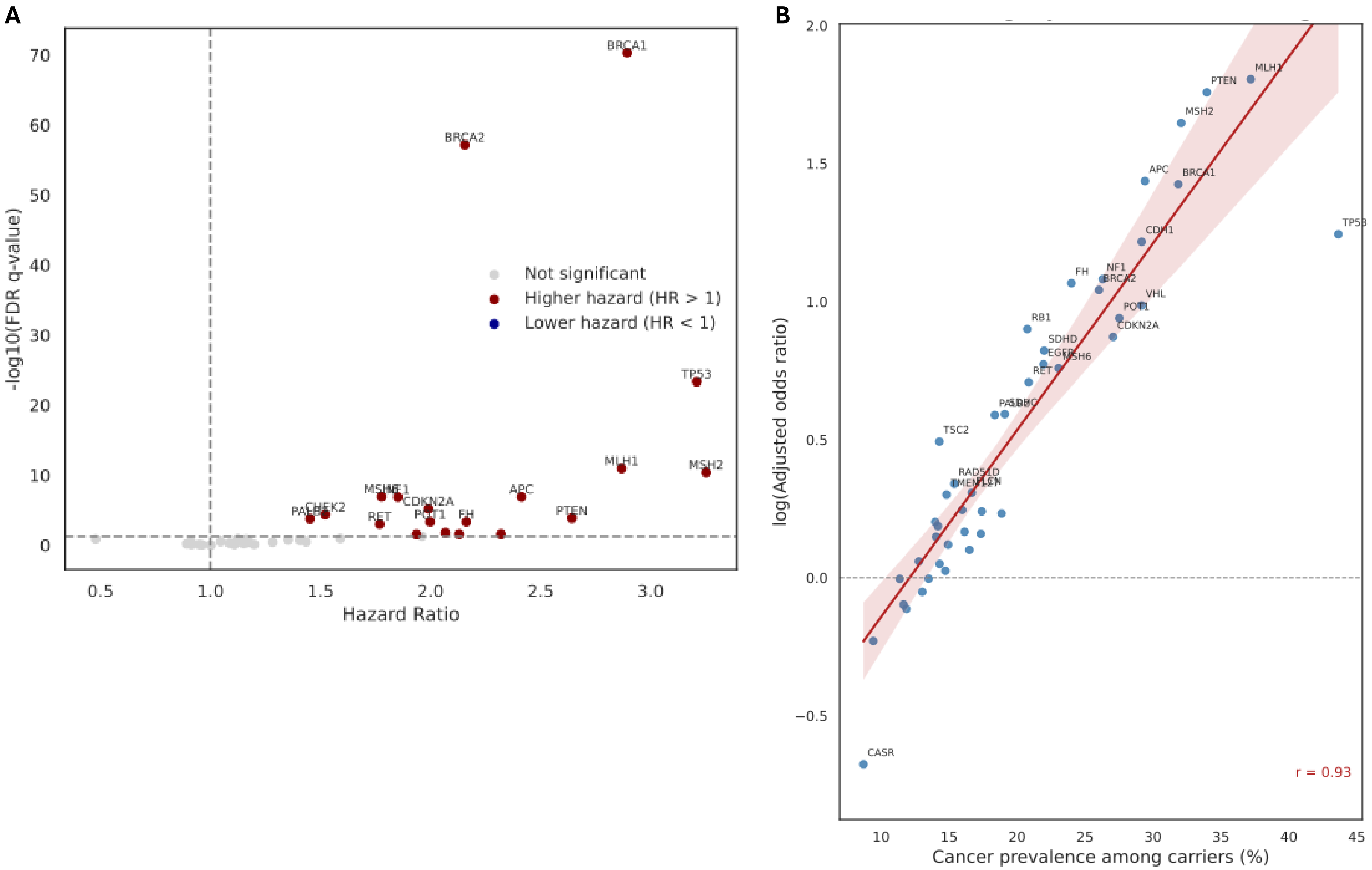

### Supplemental Figure 4

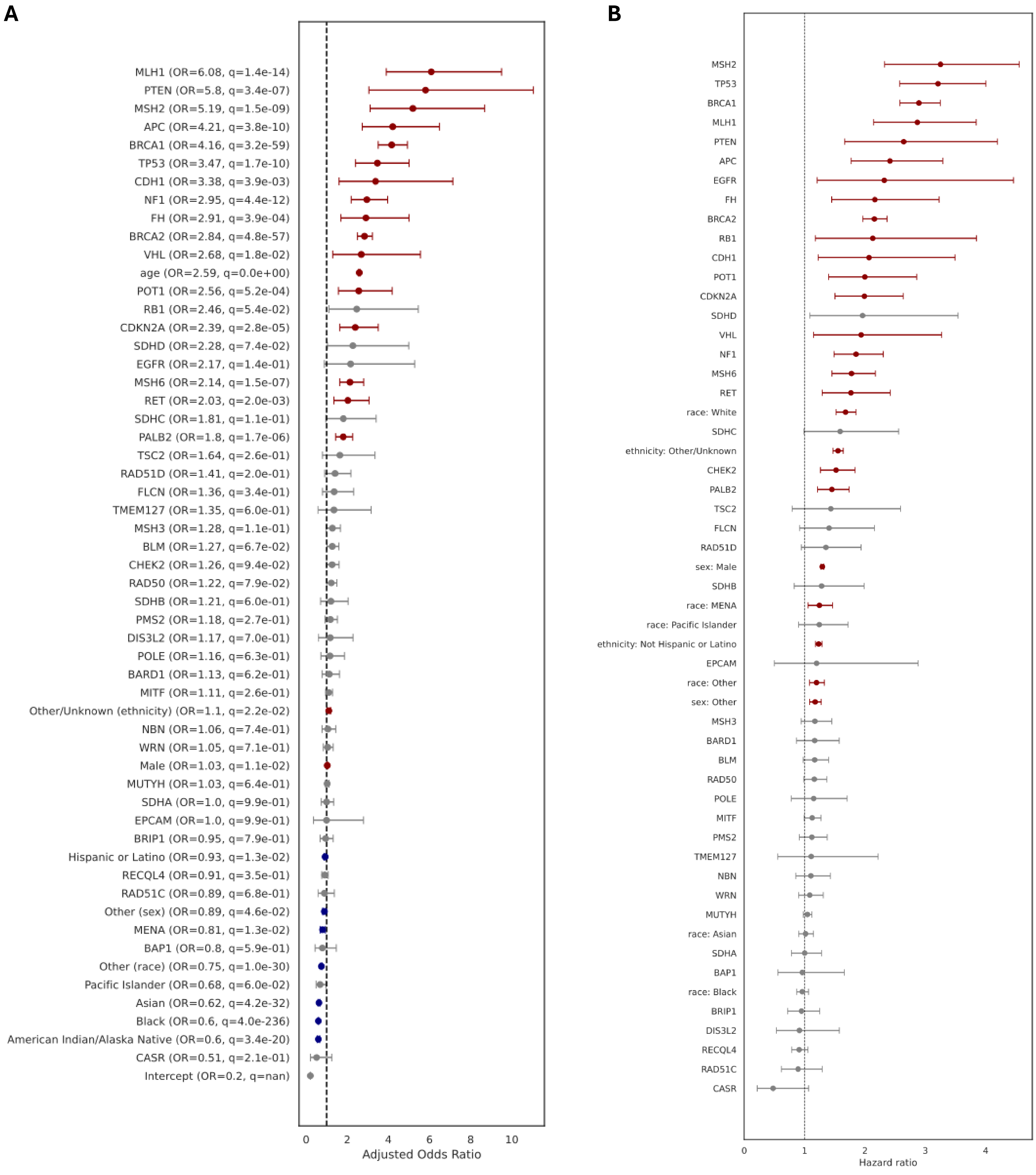

### Supplemental Figure 5

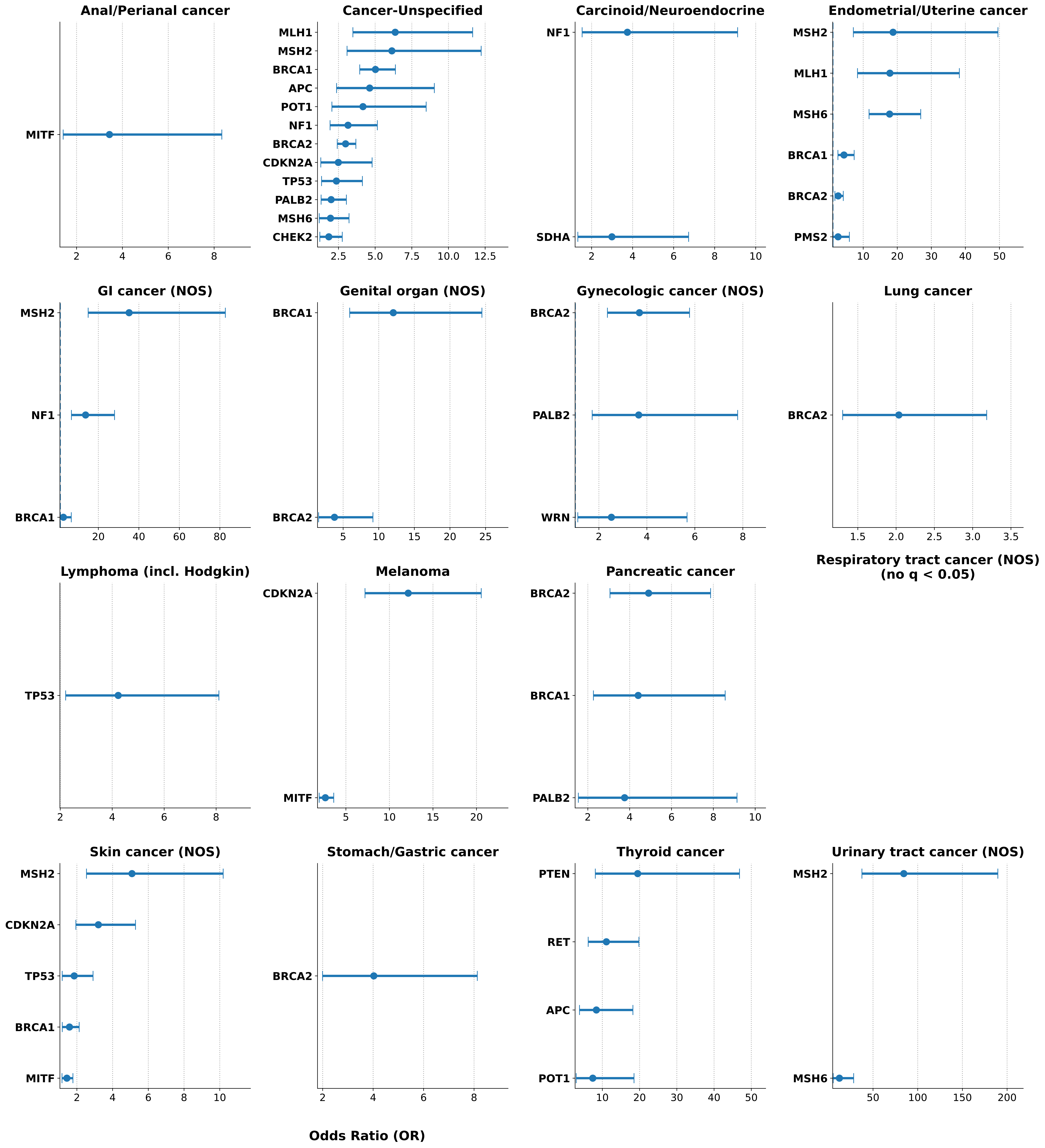
